# From Fairness Findings to Fairness Claims: An Evidence Classification Scheme for Clinical AI

**DOI:** 10.64898/2026.07.09.26357666

**Authors:** Didem Stark, Kerstin Ritter, the Alzheimer’s Disease Neuroimaging Initiative

## Abstract

Fairness audits of clinical AI models rarely make the evidentiary status of subgroup findings explicit: reassuring results may reflect insufficient statistical precision rather than true parity, and audit verdicts can easily reverse under equally defensible analytic choices. We introduce an evidence classification scheme that screens for sample size and precision, and integrates stability across design alternatives directly into the fairness claim. We demonstrate this scheme on the estimation of the brain-age gap (BAG)—a potential clinical biomarker—from structural MRI using the Alzheimer’s Disease Neuroimaging Initiative (ADNI) data. The male-female and Black-vs-White differences, along with the White-Male and Black-Female intersectional contrasts, are all classified as equivalence supported, stable across regressor choice (ridge vs. gradient-boosted trees) and feature representation (full feature set vs. cortical-thickness-only). The Asian-vs-White and Black-Male comparisons remain classified as insufficient data throughout, as neither meets the pre-specified minimum-sample threshold. The proposed scheme provides a path from raw fairness findings to justified fairness claims via pre-specified thresholds, minimum-information screening, and stability checks across declared design choices.

## 1 Introduction and Related Work

As clinical AI systems are deployed more widely, fairness audits are becoming standard practice. Yet, a reported metric is often treated as final regardless of how few patients it was based on, how common the condition was, or which design choices produced it [10]. However, small subgroups often lack the power to detect a disparity even when one exists, so a null result can reflect low power rather than true parity [26,22,7], and an observed disparity can depend on the metric or operating point used, so an equally defensible analytic choice can shrink, remove, or reverse it [14,13,21].

Several existing frameworks support fairness auditing in clinical AI. Early general-purpose toolkits such as AIF360 [3] and Fairlearn [27] compute standard group-fairness metrics across a model’s predictions; Fairlearn additionally reports bootstrap confidence intervals, cautioning that overlapping intervals do not imply statistical equality [27]. FAMEWS [8] runs early-warning systems through a multi-stage audit pipeline. Within medical imaging specifically, MEDFAIR [29] benchmarks fairness across imaging pipelines, meval [24] reports bootstrapped confidence intervals alongside its metrics, and FairLogue [23] extends the analysis to intersectional subgroups, showing that single-axis audits can understate disparities that only emerge once several attributes are considered together; separately, [16] show that a model can appear well-calibrated in aggregate while its subgroup-level calibration is poor.

A second line of work addresses the underlying statistical question directly. Cherian et al. [4] reframed auditing as a multiple-hypothesis-testing problem rather than a single pass/fail check. Ferrara et al. [7] and Singh et al. [22] address subgroup power from different angles: the former names a “no-power zone” in which subgroups are too small for any test to be informative, and the latter works out how large a subgroup would need to be to reach adequate power; Wastvedt et al. [26] show the same problem concretely in small racial subgroups, where a handful of observations in a key outcome cell can widen confidence intervals past the point of usefulness. Metric choice adds a further complication: McDermott et al. [13] show that AUPRC can systematically favor whichever subgroup is more prevalent, and Simson et al. [21] demonstrate that a fairness verdict can be pushed almost anywhere across a defensible “multiverse” of design choices. Equivalence and non-inferiority testing offers one way to address this gap: Schuirmann’s Two One-Sided Tests procedure [20] is the standard approach in pharmacology, Lakens [11] brought it into psychology, and it has begun to appear in fairness testing as well, in work examining pricing algorithms [9] and general fairness-testing frameworks [12].

The proposed scheme first screens whether the data are precise enough to say anything about a pre-specified margin, then applies a Two One-Sided Tests (TOST) equivalence procedure against that margin, and finally checks whether the conclusion holds under reasonable changes in model choice or margin size.

This yields a claim-specific evidence state for each subgroup — *insufficient data, imprecise, equivalence supported*, or *equivalence not supported* — each further marked *stable* or *conditional* depending on whether it holds up across analytic choices. We demonstrate the classification scheme on the estimation of the age-debiased brain-age gap (BAG) from sMRI in ADNI. BAG is a potential clinical marker associated with mortality and age-related health outcomes [5,6,28], so auditing parity in its estimation across groups is critical: performance differences here could translate into systematic over- or under-diagnosis of age-related cognitive decline or neurodegenerative disease in underrepresented groups. Most sex-fairness studies in brain age prediction from sMRI rely on significance testing without a pre-specified margin [17,18]. Race-based analyses are rarer, despite well-documented enrollment imbalances in cohorts such as ADNI [2]; where reported, Black-White brain-age gap disparities are around 1–2 years and vary by algorithm [17,1]. Our scheme separates findings that are stable across model and feature choice from subgroups that remain genuinely unresolved due to insufficient data.

## 2 Evidence Classification Scheme

The proposed scheme treats fairness auditing as a sequential process (Figure 1), requiring the prior specification of subgroup metrics, a reference group and a target margin.

**Fig. 1.**
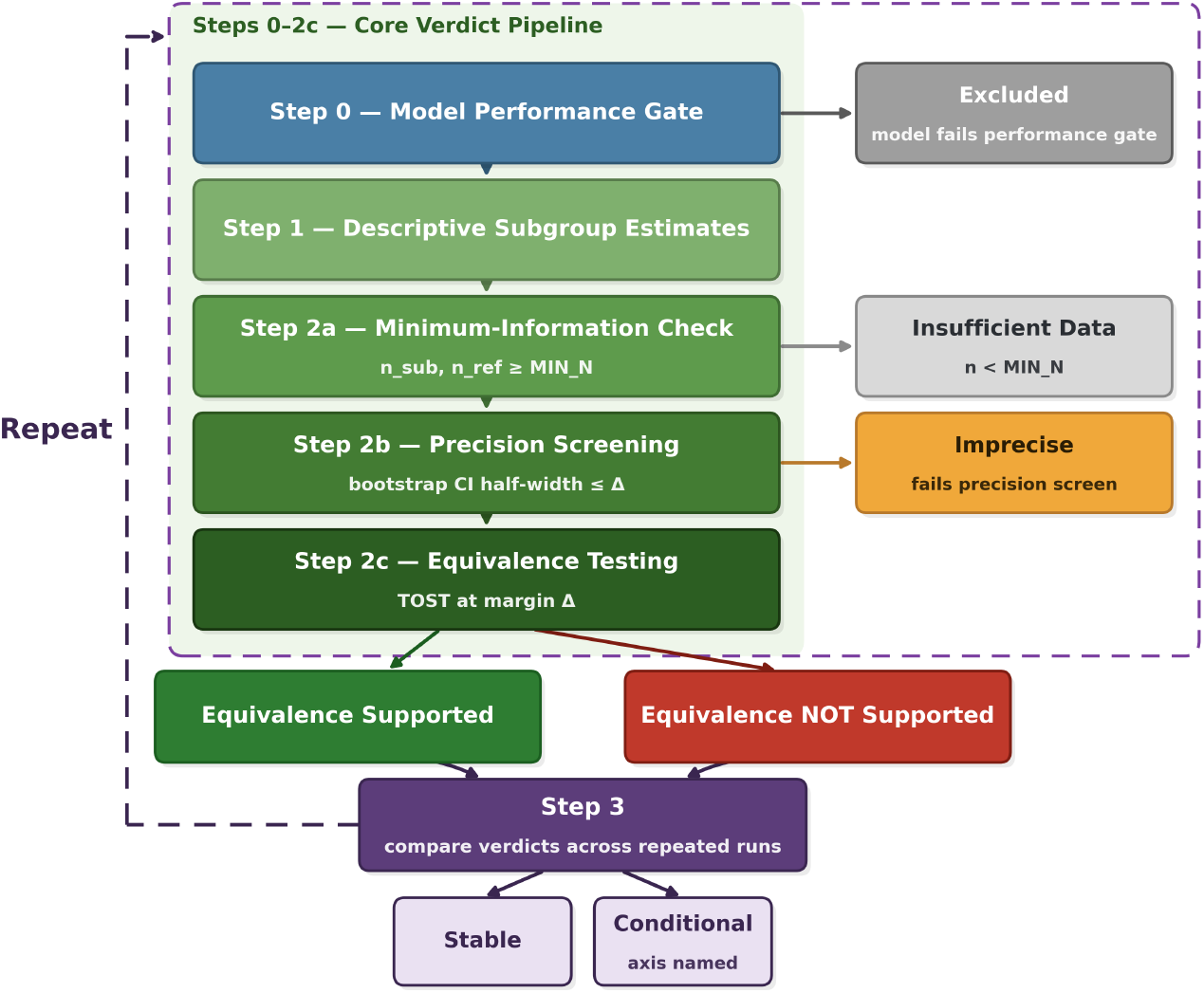
Evidence classification scheme for subgroup fairness claims, from performance and precision checks to equivalence inference and stability assessment.

### Step 0 — Performance gate

The model must meet pre-specified, task- and stakeholder-defined minimum performance thresholds. A model failing this gate should not be advanced as deployment-ready, and any subgroup audit becomes diagnostic only.

### Step 1 — Initial finding

Records the observed subgroup metric pattern, using a task- and stakeholder-defined metric, which may be either a standard performance metric (e.g. AUROC or false negative rate) or a common fairness metric (e.g. equalized odds or equal opportunity). Both apparent fairness and apparent disparity require further validation before any conclusions can be drawn.

### Step 2a — Minimum-information check

Subgroups below a minimum sample size are classified as *insufficient data* without proceeding to further testing.

### Step 2b — Precision screening

This step checks whether the data are sufficiently precise to draw conclusions relative to the specified margin. Let the symmetric bootstrap half-width (HW) be half of the confidence interval (CI) obtained by bootstrapping: HW = (CI_hi_ *−* CI_lo_)*/*2; subgroups with HW *> Δ* are classified as *imprecise*.

### Step 2c — Equivalence testing

Subgroups that pass precision screening are evaluated using the Two One-Sided Tests (TOST) procedure [20,11]. Let *d* = *m*_subgroup_ *−m*_ref_ denote the subgroup–reference contrast and *Δ >* 0 the target margin: the TOST null hypothesis *H*_0_ : |*d*| ≥*Δ* is rejected — yielding *equivalence supported* — when the confidence interval lies entirely within (*−Δ*, +*Δ*), and *equivalence not supported* otherwise. We report a two-sided 95% confidence interval throughout, corresponding to *α* = 0.025 per tail. Note that because signed parity reflects calibration relative to a reference group, an equivalence verdict remains distinct from absolute calibration against zero error.

### Step 3 — Stability to design choices

This step evaluates verdict stability across pre-specified alternatives, such as different model families or feature sets. Verdicts unchanged across all alternatives are labeled *stable*; verdicts that change are labeled *conditional*, with the responsible axis identified explicitly — for example, *conditional on model family*.

## 3 Case Study: Brain-Age Gap Estimation

### 3.1 Data, Cohort Construction, and Preprocessing

We validate the scheme using sMRI data from stable cognitively normal (CN) participants across phases 1/GO/2/3/4 of the ADNI (adni.loni.usc.edu), using the UCSF FreeSurfer cross-sectional table `(UCSFFSX7)`, demographics `(PTDEMOG)`, and diagnostic summary (DXSUM) (tables extracted 10 June 2026). We define stable CN status as ever receiving a diagnosis code of CN (1) and never receiving a code of MCI (2) or AD (3) at any visit. We restrict to 3T acquisitions and filter intracranial volume (ICV, FreeSurfer field ST10CV) to fall within [500,000, 2,000,000] mm^3^; additionally check for `(OVERALLQC = Pass)` in ADNI4. We restrict subjects to recorded sex codes (male/female), compute chronological age as scan year minus birth year and restrict it to [50, 100], and drop duplicate RID–exam-date rows, keeping the first. We derive race from PTRACCAT, collapsed to White, Black, Asian, or Other (all remaining codes).

We use a subject-level split stratified jointly by sex and race: within each sex×race stratum, ⌊*n/*2⌋ subjects are randomly assigned to the held-out test set, with the remainder forming the cross-validation (CV) pool; strata with fewer than 2 subjects are excluded from splitting. The test set retains one scan per subject (earliest exam date). Resulting cohort size, demographic composition, and split sizes are reported in Table 1.

**Table 1.**
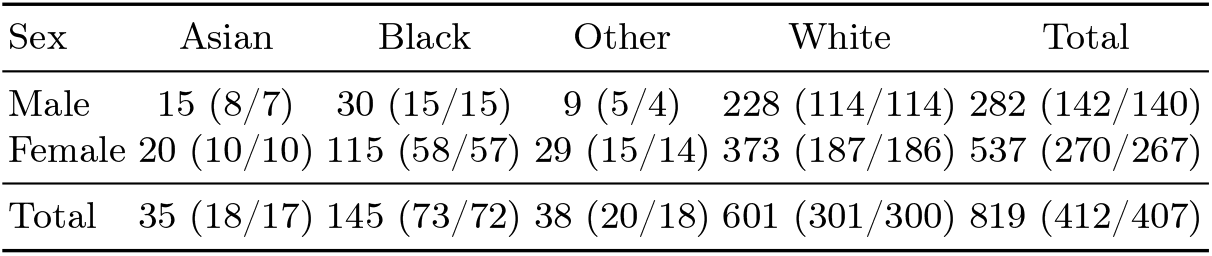
Cohort demographics by sex and race, at the subject level. Each cell gives *N* (Train/Test). Race category “Other” is excluded from race-based contrasts (Table 2) but included in row/column totals.

FreeSurfer structural features comprise all cortical and subcortical volumes, surface-area and cortical thickness values. Volume and surface-area features (`*CV` cortical volume, `*SV` subcortical volume, `*SA` surface area) are residualized against ICV to remove head-size scaling: for each such feature with more than 20 non-missing training observations, we estimate a linear fit (feature∼ ICV) on the training partition only, and correct both training and test values as 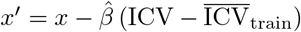. Cortical-thickness features (`*TA`, cortical thickness average, `*TS` cortical thickness standard deviation) are left uncorrected. We drop features with more than 10% missingness in the training partition; the remainder are median-imputed and standardized (z-scored), with both the imputer and scaler fit exclusively on the training partition and applied unchanged to the test partition.

### 3.2 Model, Audit Configuration, and Stability

We fit a ridge regression model to predict chronological age from the preprocessed features, with the regularization strength selected via 5-fold subject-grouped cross-validation (GroupKFold) over *α* ∈{0.01, 0.1, 1, 10, 100, 1000, 10000}, minimizing mean squared error; the selected value is reported in Section 4. Additionally, we apply age-bias correction following [5,6]: a linear model of prediction error as a function of chronological age is fit on the training partition’s out-of-fold CV errors, and this trend is subtracted from the test-set errors.

We illustrate the audit with the following parameters: The Step 0 gate requires the CV pool Mean Absolute Error (MAE) to fall below 80% of a dummy regressor predicting the training mean age; this gate is evaluated on raw (uncorrected) predictions, as it screens the model’s basic ability to learn an age-relevant signal, independent of how that signal is subsequently expressed as a clinical marker. The minimum-information check (Step 2a) requires *n* ≥ 20 per subgroup, evaluated on the held-out test-set count for that subgroup (Table 1), since all reported contrasts and CIs are computed on test-set predictions. We test the signed mean-error difference: 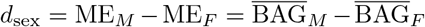 (reference: female) and 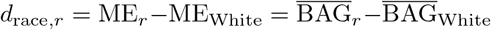 for *r* {Black, Asian} (reference: White; race category Other is excluded from race-based contrasts); intersectional subgroups (White Male, Black Female, Black Male) are compared against a White Female reference. We take female as the sex reference following prior subgroup analyses of brain-age prediction, which found male subjects to be the more negatively affected group in model performance [17]; White is retained as the race reference given its majority representation in ADNI [2].

*p*-values from the bootstrap TOST-style test (Section 2), each based on a two-sided 95% CI (*α* = 0.025 per tail), are additionally Benjamini–Hochberg adjusted at a family-wise *α* = 0.05 across all contrasts with sufficient data. CIs are percentile bootstraps over 1,000 subject-level resamples, drawn independently with replacement within the subgroup and reference sets (stratified by the compared subgroups), using a reproducible seed per contrast. Comparisons used an illustrative target margin of *Δ* = 2 years, based on the 1–2 year disparity range previously reported across models [1] and consistent with per-year mortality-risk associations reported for the brain-predicted age gap biomarker [5].

Stability is assessed along two axes, both under the same train/test split: (i) *model*, comparing the primary ridge model against a gradient-boosted tree model (XGBoost) on the full feature set, with hyperparameters selected from a fixed grid of four conservative configurations via the same Group KFold procedure used for ridge; and (ii) *features*, comparing the primary full-feature ridge model against a ridge model restricted to cortical-thickness features only, as an alternative feature set. A model × feature combination (XGBoost, thickness-only) varying both axes at once is also run as a confirmatory check but is not independently scored. A contrast is labeled *stable* if its classification is unchanged across all scored arms, and *conditional (axis)* otherwise, naming the axis (or axes) responsible for the change.

## 4 Results

### 4.1 Cohort, Performance Gate, and Main Audit

The cohort comprised 819 subjects (2,121 scans), split at the subject level, stratified jointly by sex and race, into a training/CV pool of 412 subjects (1,062 scans) and a held-out test set of 407 subjects (407 scans, one scan per subject). Table 1 provides the full breakdown by sex and race.

The primary ridge model (GroupKFold-selected *α* = 1000, 321 retained features) passed the Step 0 gate: out-of-fold CV MAE = 4.24 against a dummy-predictor MAE of 5.76 (threshold 4.61); the Cole-corrected CV MAE was lower still, at 2.83, also passing. Cole-corrected test-set MAE was 2.69, corresponding to a mean absolute BAG of 2.69 years on the held-out set, and forms the basis for all subgroup contrasts reported below.

Table 2 reports the six pre-specified contrasts at *Δ* = 2 years under the primary model audited. Asian-White and Black-Male fail the minimum-information check (*n* ≥ 20 on the held-out test set; Table 1) and are classified as insufficient data. The remaining four contrasts all pass precision screening (Step 2b) and are subsequently classified as *equivalence supported* under equivalence testing (Step 2c): each CI is fully contained within (−2, 2), and each TOST test remains significant after Benjamini–Hochberg correction across the four-contrast family (*p*_*adj*_ = 0.001 throughout).

**Table 2.**
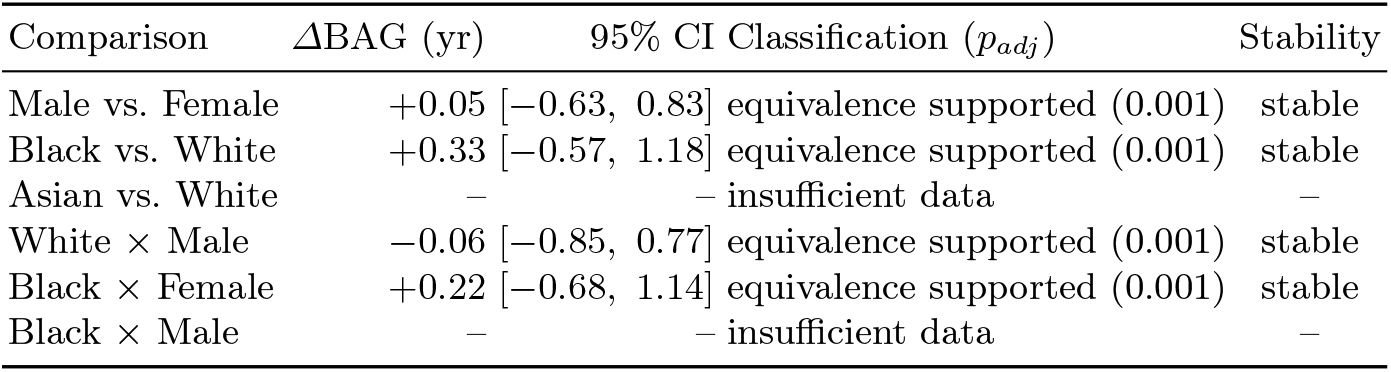
Step 1–2 classification at *Δ* = 2 years, with Step 3 Stability verdicts. *Δ*BAG is the signed BAG contrast (subgroup*−* reference, years), as defined in Section 3.2; CI is the 95% bootstrap interval; TOST *p* is BH-corrected across the four testable contrasts and floored at 1*/*1,000 by the bootstrap resolution, which all four contrasts reach here. Stable reflects agreement across model choice (ridge/XGBoost) and feature set (full/thickness-only) (Section 3.2).

### 4.2 Stability

All four testable contrasts are classified identically under both scored Step 3 axes: switching the model from ridge to XGBoost (hyperparameters {300 estimators, max depth 3, learning rate 0.02, subsample 0.6, column-subsample 0.4, *ℓ*_2_ weight 10}, selected by the same GroupKFold procedure), and switching the feature set from the full feature set to a 136-feature cortical-thickness-only set (which bypasses the ICV correction entirely), each independently clearing the Step 0 gate (CV MAE 4.26 and 4.52 respectively, against the 4.61 threshold). All four contrasts are therefore labeled *stable*; the confirmatory XGBoost+thickness-only arm failed the Step 0 gate on raw CV MAE (4.66 *>* 4.61).

## 5 Discussion

In this work, we propose an evidence classification scheme for interpreting subgroup fairness findings in clinical AI, separating insufficient-data and imprecise comparisons from margin-based verdicts qualified by stability across declared design alternatives. A fairness audit report should state whether its evidence is insufficient, conditional, or stable within a declared design space. We then demonstrate it on BAG prediction from MRI on ADNI. Under the primary configuration (Table 2), all four testable contrasts are classified as equivalence supported and remain so under both model architecture and feature representation. The minimum-information check likewise proves consequential: Asian-White and

Black-Male both fall below the *n* ≥ 20 threshold and are classified as insufficient data. Instead of reporting an underpowered estimate that could be mistaken for either reassurance or evidence of disparity, the scheme declines to test these contrasts at all, which is a deliberate design choice rather than a limitation of the bootstrap procedure. These results illustrate why clinical fairness audits should not be reduced to a single binary verdict, even when metric, cohort, and model specification are held fixed.

Beyond the audit itself, this distinction bears on medical device regulation and algorithmic governance, which increasingly demand auditable evidence of parity rather than a stated principle [15,25]. A margin-backed, state-qualified verdict such as insufficient data, stable, or conditional on a named design choice offers stakeholders a more informative vocabulary than a raw performance gap or a single pass/fail outcome [19].

BAG is associated with relative risk profiles for neurodegeneration, cognitive impairment, and all-cause mortality [5,6,28], so understanding systemic demographic imbalances in its derivation is crucial. An algorithm that systematically over- or underestimates a clinical score for a given demographic group introduces bias into downstream diagnostic logic: a *conditional* or *unsupported* equivalence verdict, if overlooked, could permit the systematic over-diagnosis of pathological cognitive decline in some subgroups, or the under-diagnosis of neurodegenerative disease in others, with correspondingly unequal clinical consequences.

Several limitations apply. These results are specific to the ADNI cohort’s composition, its recorded sex and race categories, and the regression architectures used, drawing on FreeSurfer 7 features. The illustrative *Δ* = 2-year margin requires validation against concrete clinical endpoints. The case study is retrospective and illustrative: the scheme requires subgroup definitions, metrics, reference groups, margins, tests, and design alternatives to be declared prospectively.

More broadly, the scheme does not resolve which reference group or disparity threshold is appropriate; it renders that choice visible and auditable. Furthermore, the framework currently relies on an arbitrary minimum sample size (*n*) threshold to filter out small cohorts, which risks either prematurely excluding minoritized groups or reporting volatile, noisy metrics; however, this choice can be mathematically formalized in production using methods such as statistical power analysis. Finally, defining a performance margin that is truly anchored to concrete clinical endpoints remains a core challenge, as translating mathematical algorithmic variance into predictable, real-world impacts on patient outcomes requires extensive empirical validation.

## Data Availability

All data used are available at ADNI (https://adni.loni.usc.edu/).

## Acknowledgments

This research was funded by the Deutsche Forschungsgemeinschaft (DFG) through the Walter Benjamin Programme (project number 565037378). Additional support was provided by the Gemeinnützige Hertie-Stiftung, the Tübingen AI Center, and the German Center for Mental Health (DZPG).

Data used in preparation of this article were obtained from the ADNI database (https://adni.loni.usc.edu/). ADNI investigators contributed to the design and implementation of ADNI but did not participate in analysis or writing of this report. A complete listing of ADNI investigators can be found at:http://adni.loni.usc.edu/wp-content/uploads/how_to_apply/ADNI_Acknowledgement_List.pdf.

Generative AI was used for the preparation of this manuscript.

The authors have no competing interests to declare.

## Notes

### Competing Interest Statement

The authors have declared no competing interest.

### Author Declarations

Alzheimer's Disease Neuroimaging Initiative (ADNI) https://adni.loni.usc.edu/

